# Investigating sodium homeostasis of structural brain hubs in focal epilepsy using 7T MRI

**DOI:** 10.1101/2025.01.02.25319878

**Authors:** Lucas Gauer, Roy AM Haast, Mikhael Azilinon, Mohamed Mounir El Mendili, Julia Scholly, Hugo Dary, Vera Dinkelacker, Jean-Philippe Ranjeva, Wafaa Zaaraoui, Fabrice Bartolomei, Maxime Guye

## Abstract

Besides their crucial role in cerebral connectivity, brain hubs are regions vulnerable to the energy deficit associated with various brain disorders. Changes in sodium homeostasis of cortical regions have been observed in focal epilepsy and may reflect energy failure. We investigated whether nodal structural connectivity is differently affected within the hub and non-hub regions by ionic perturbations associated with focal epilepsy. Our hypothesis was that the metabolic demands of hub regions may be associated with a distinct ionic profile detectable by sodium MRI and that this profile is altered in focal epilepsy.

We recruited 23 patients with drug-resistant focal epilepsy and 21 age- and gender-matched healthy controls. Anatomical, diffusion-weighted, and sodium imaging was performed using a 7 Tesla MRI scanner. Patients underwent pre-surgical work-up, including stereo-electroencephalographic recordings for defining the epileptogenic regions. Anatomical parcellation and multimodal coregistration allowed the use of parcels as nodes of whole-brain structural connectomes, linking structural connectivity measures to epileptogenicity and sodium parameters. Sodium parameters in patients were z-scored concerning homologous parcels in controls to allow comparison across regions of interest.

Hub regions had higher total sodium concentration (TSC) than non-hub regions in both patients and controls, and this difference was not observed for sodium signal fraction (*f*, a proxy of intracellular sodium homeostasis). Compared to controls, patients showed increased TSC in both epileptogenic and non-epileptogenic zones, and this increase in TSC was consistent in both hub and non-hub regions. On the opposite, *f* was increased only within the epileptogenic zones and was not affected by the hubness of a region.

Our results confirm the whole brain increase in TSC and the local increase of the *f* value within epileptogenic zones previously observed in focal epilepsy patients. Therefore, we propose that that sodium imaging can probe distinct tissue properties: TSC appears sensitive to microstructural alterations, while *f* could reflect homeostatic disruptions specific to epileptogenic regions.

## Introduction

Focal epilepsy occurs when abnormal rhythms arise within a network of anatomically and functionally connected brain regions (*i.e.*, the epileptogenic zone, EZ). These rhythms can further propagate to other regions connected to this epileptogenic network (*i.e.*, the propagation zone, PZ), leading to the clinical symptoms observed during a seizure.^1–4^ Abnormal rhythms during seizures are typically understood as patterns of sustained rhythmic depolarization of neurons and the resulting electrical activity recorded by scalp EEG or intracranial recordings like stereoelectroencephalography (SEEG). However, these neuronal firing rhythms are just one example of the broader range of rhythmic processes that occur in the brain in physiological or pathological conditions. Indeed, seizures also disrupt a range of other rhythmic activities occurring at different biological scales, such as the dynamic equilibrium (homeostasis) of ion and molecule exchanges between neurons and glial cells, energy consumption rates, local blood supply, and waste product clearance.^5–14^

Techniques that probe the multifactorial disruption of homeostasis are pivotal to provide a broader picture of focal epilepsy and its consequences. Unlike conventional proton-based (^1^H) MRI, sodium (^23^Na) MRI can detect ionic changes associated with energy failure in various brain disorders.^15–20^ In focal epilepsy, this technique showed a global increase in cortical total sodium concentration (TSC).^21^ Furthermore, using ultra-high field ^23^Na MRI at 7 Tesla (7T), the (effective) transverse (T_2_*) relaxation of the sodium signal can be recorded and modeled as a biexponential function with a fast (T_2_*_short_) and a slow (T_2_*_long_) decay factor. In a previous study, the ratio (*f*), which is the ratio of sodium signal from the fast component and TSC, was shown to increase within the EZ but not in the PZ or non-involved zones (NIZ) compared to controls, demonstrating that ^23^Na MRI can be sensitive to sodium homeostasis alterations specific to regions characterized by high epileptogenicity.^22^

Beyond the epileptic networks, the brain can be viewed as an extensive network itself. In this representation, nodes are defined by anatomical regions, and edges by the connections between pairs of nodes, collectively forming a connectome. These connections can be based on number of fiber bundles connecting identified from diffusion MRI data (structural connectome) or the temporal correlation of the blood oxygen-level dependent signal in functional MRI data (functional connectome).^23,24^ Applying graph metrics to these connectomes allows us for the characterization of certain regions (or nodes) as hubs of structural or functional connectivity, respectively.^25,26^ Studies have shown that hub regions are often at the epicenter of various neurological diseases.^27^ One hypothesis is that the high metabolic demands of hub regions—often referred to as “wiring cost”—could make them more vulnerable to energy deficits and homeostatic alterations associated with pathological states.^28–30^

The relationship between hubs and epilepsy, however, remains incompletely understood. Studies suggest that patients with more topologically isolated epileptogenic zones or those who had hubs resected tend to have better surgical outcomes. In contrast, centrally integrated epileptogenic regions are associated with higher recurrence rates.^31,32^ Increased global efficiency of information flow has been linked to a greater risk of postoperative seizures, while reduced connectivity between regions where interictal epileptic discharges occur and other cortical areas has been associated with seizure freedom.^33,34^ Furthermore, approaches using structural connectome data and hub metrics have successfully predicted seizure outcomes, emphasizing the critical role of network configuration and specific hubs, such as the thalamus, in seizure control and cognitive function.^35^ These findings underscore the importance of hub integrity in surgical outcomes and suggest that further investigation into hub physiology in epilepsy could aid in refining therapeutic strategies.

Building on the aforementioned findings and addressing the current knowledge gap regarding hub integrity, this study aims to investigate whether hub and non-hub nodes are similarly affected by the ionic perturbations associated with focal epilepsy. We hypothesize that the high metabolic demands of hub regions may be associated with an unique ionic profile, which can be studied using ^23^Na MRI and that this profile may be altered in the context of focal epilepsy.

## Materials and methods

### Subjects

We included patients with drug-resistant focal epilepsy (DRFE) undergoing presurgical evaluation and requiring SEEG at La Timone University Hospital in Marseille between June 2017 and November 2020. For these patients, SEEG was needed when the precise delineation of the EZ was not achieved by a previous non-invasive exploration (*i.e.,* combining long-term video-EEG monitoring, morphological MRI, [^18^F]fluorodeoxyglucose positron emission tomography and/or magnetoencephalography). Based on these non-invasive investigations, SEEG implantation was planned for each patient according to anatomo-electro-clinical hypotheses about the localization of the EZ network.^36^ All patients underwent a high-resolution 7T MRI protocol combining anatomical ^1^H MRI, diffusion-weighted imaging (DWI), and ^23^Na MRI before SEEG implantation (see below). We also used an age and sex-matching control database of 23 healthy subjects who underwent the same 7T MRI protocol.

From our initial cohort of 64 patients and 23 healthy subjects, we included 23 patients (mean age 32 ± 11 years, 50 % female) and 21 healthy controls (mean age 38 ± 16 years, 57 % female). We excluded 19 patients with major anatomical brain defects that would interfere with automatic segmentation *(i.e.,* major hemispheric vascular malformations, porencephalic cavity, extensive cortical malformations) from further analysis within the scope of this study. We also excluded 16 patients who did not undergo SEEG, two patients in whom no seizure was recorded during the SEEG procedure, and four patients after image quality check in the three modalities (anatomical, DWI, and ^23^Na MRI). Finally, poor image quality led to the exclusion of 2 healthy controls after the image quality check (Supplementary Fig. 1).

### MRI acquisition protocol

Participants of this study (EPINOV trial, NCT03643016 and the local EPI study) provided informed consent in compliance with the ethical requirements of the Declaration of Helsinki and the protocol was approved by the local Ethics Committee (*Comité de Protection des Personnes sud Méditerranée 1*). This study complies with the requirements of the national personal data regulation committee (*Commission Nationale de l’Informatique et des Libertés*).

For each subject, images were acquired during one session, using a whole-body 7T Magnetom Step 2 MR system (Siemens Healthineers, Erlangen, Germany). Anatomical and DWI ^1^H imaging was acquired using a 1Tx/32Rx ^1^H head coil (Nova Medical, Wilmington, USA). Sodium imaging was conducted with a dual-tuned ^23^Na/^1^H QED birdcage coil.

Anatomical images consisted of 3D Magnetization-prepared two rapid gradient-echo (MP2RAGE) (TR = 5000 ms, TE = 3 ms, TI1 = 900 ms, TI2 = 2750 ms, 256 slices, 0.6 mm isotropic resolution, acquisition time = 10 min) acquired after an automatic B_0_ shimming procedure and a whole brain B_1_^+^ map derived from a spin-echo-based sequence, by assessing the ratio of consecutive spin and stimulated echoes (WIP#658, Siemens Healthineers, Erlangen, Germany).

Diffusion imaging was conducted using two ^1^H-MRI DWI acquisitions (1.12 mm^3^, 80 directions, b-values = 1000 and 2000 s/mm^−2^, with interleaved b_0_) with two opposite phase-encoding directions (acquisition time = 11 min for each phase encoding direction).

Sodium MRI was conducted using a protocol described previously.^22^ Briefly, we used a multi-echo density adapted 3D projection reconstruction pulse sequence (TR = 120 ms, 5000 spokes, 384 radial samples per spoke, 3 mm nominal isotropic resolution, 24 echoes times from 0.20 to 70.78 ms, acquisition time = 30 min in total).^37^ For quantitative calibration of brain sodium concentrations, we used as external reference six tubes (80 mm length, 15 mm diameter) positioned in the field of view in front of the subject’s head and filled with a mixture of 2 % agar gel and sodium at different concentrations (2x25 mM, 50 mM, 2x75 mM, and 100 mM).

### Anatomical MRI processing

Automatic segmentation and parcellation were performed on the B_1_^+^-corrected T_1_w (i.e., ‘UNI’) MP2RAGE images using FreeSurfer (v6.0, available at https://surfer.nmr.mgh.harvard.edu) and the Virtual Epileptic Patient (VEP) atlas. This led to the identification of 81 regions per hemisphere, including cortical, subcortical regions and cerebellar hemispheres.^38,39^

### Diffusion MRI processing

DWI images were denoised and corrected for Gibbs artefacts and image bias using MRtrix3 tools.^40^ Spatial distortions introduced by echo-planar imaging were corrected using the interleaved b_0_ images with opposite phase-encoding directions using FSL’s *topup* tool. Eddy currents were corrected using the *eddy* tool (available at http://fsl.fmrib.ox.ac.uk). Anatomical MP2RAGE were coregistered to DWI images (ANTs v4.9^41^) and 10 million streamlines were generated using anatomically constrained tractography.^40,42^ Raw tractograms were filtered to 1 million streamlines using SIFT algorithm.^43^ Weighted matrices of the connectomes were generated for each subject using the VEP region as nodes and streamlines count between each pair of regions as edge weights (parameters in Supplementary File 1).

### Sodium MRI processing

Multi-echo sodium images were processed using a method previously described using the relevant functionalities implemented in SPM12.^21,18,22^ Briefly, multi-echo sodium images were denoised with a rician filter and corrected for motion.^44,45^ T_1_w images were coregistered to the sodium images and the associated VEP atlas parcellation. T_1_w images were segmented to generate a CSF probability map. Voxels with a probability of CSF > 0.1 were excluded from the parcels, and the atlas was resampled into the sodium space using the nearest neighbor interpolation. The mean sodium signal within each parcel was then fitted using custom MATLAB code with a bi-exponential model and normalized relative to signals from reference tubes to compute the mean total sodium concentration (TSC) and sodium signal fraction (*f*) as previously described.^22,37^

### SEEG recordings

Recordings were performed using intracerebral multi-contact electrodes (10-18 contacts, length 2 mm, diameter 0.8 mm, 1.5 mm apart, Alcis, France). Electrodes were implanted based on the clinical hypotheses of the EZ and seizure propagation pathways using a ROSA™ stereotactic surgical robot. A cranial CT scan was performed to verify the absence of complications and the spatial accuracy of the implantation. The signals were recorded on a 256-channel Natus system, sampled at 512 Hz, and stored on a hard disk (16 bits/sample) without digital filtering. Two hardware filters were present in the acquisition procedure: a high-pass filter (cut-off frequency equal to 0.16 Hz at -3 dB) and an anti-aliasing low-pass filter (cut-off frequency equal to 170 Hz at 512 Hz).

### SEEG signal analysis

For each patient, epochs of recorded seizures were selected for analysis, and the signal from contacts located in the grey matter was analyzed using AnyWave software.^46^

The signal of each contact was visually checked by an expert (JM), and contacts with artifacts were excluded from the analysis. The Epileptogenicity Index (EI) of each bipolar derivation was computed.^47^ Based on previous studies, an EI value of 0.4 and higher was used to define a bipolar derivation as belonging to the EZ. In contrast, the PZ was defined as derivations with 0.1 < EI <0.4, with sustained discharge during the seizure.^22,48,49^ We labeled each pair of bipolar SEEG contacts as belonging to the EZ, PZ, or NIZ, as defined by the EI value and after visual inspection (JM) of each seizure to ensure the clinical and electrophysiological validity of the results.

By co-registering the post-implantation cranial CT scan with the pre-implantation MRI scan using Gardel software^50^, each bipolar derivation was localized within the patient’s VEP atlas parcellation. Each parcel could thus be defined as belonging to the epileptogenic zone (EZ), the propagation zone (PZ), the non-involved zone (NIZ) or the unexplored zone.

### Graph analysis

For each subject connectome, network measures were computed using *bctpy* (https://github.com/aestrivex/bctpy), the Python version of the Brain Connectivity Toolbox.^51^ The following measures were extracted: parcel’s degree (and strength), betweenness centrality, clustering coefficient, average path length. A hub score combining these measures was calculated according to Van den Heuvel et al.^52^ In short, this score represents the number of times that a node belonged to the 20% of nodes presenting the highest strength, highest betweenness centrality, lowest clustering coefficient and/or lowest characteristic path length. Parcels with a hub score ≥ 2 were defined as hubs, as previously described.^52^

### Statistical analysis

Robust z-scores (using the median and scaled median absolute deviation) were chosen over traditional z-scores (based on the mean and standard deviation) to ensure validity when applied to non-normal or skewed distributions with outliers.^53^ Regions of interest were defined as ensembles of parcels sharing the same label: (i) hub or non-hub status(ii) or EZ, PZ, NIZ, or non-explored label. The z-score distributions across regions of interest were compared using a non-parametric permutation test with 10,000 iterations. Bootstrap samples were generated by resampling (with replacement) from patient and control z-score distributions. For each iteration, the mean difference between patient and control samples was calculated. A 95 % confidence interval for the mean difference was determined using the 2.5^th^ and 97.5^th^ percentiles of the bootstrap differences. P-values were computed by comparing the observed mean difference to the bootstrap distribution. To correct for multiple comparisons, we applied a false discovery rate (FDR) adjustment with a statistical significance threshold of 0.05.^54^

### Graphical summary

A graphical summary of the methods used is shown in Supplementary Fig. 2.

### Data availability

The data supporting the findings of this study are available from the corresponding author upon reasonable request.

## Results

### Patient characteristics

The clinical characteristics of the patients are described in Table 1. The majority of patients had MRI-negative epilepsy (73 %), and had an epileptic network involving the temporal lobe (65 %, Figure 1). The mean duration of epilepsy in the entire cohort was 17 years. All patients with a lesion visible on the MRI were seizure-free (Engel class I) at least one year after surgery, while 75 % of the patients with MRI-negative epilepsy were either seizure-free (Engel class I: 37,5 %) or had only limited seizures (Engel class II: 37,5 %) after surgery.

**Figure 1:**
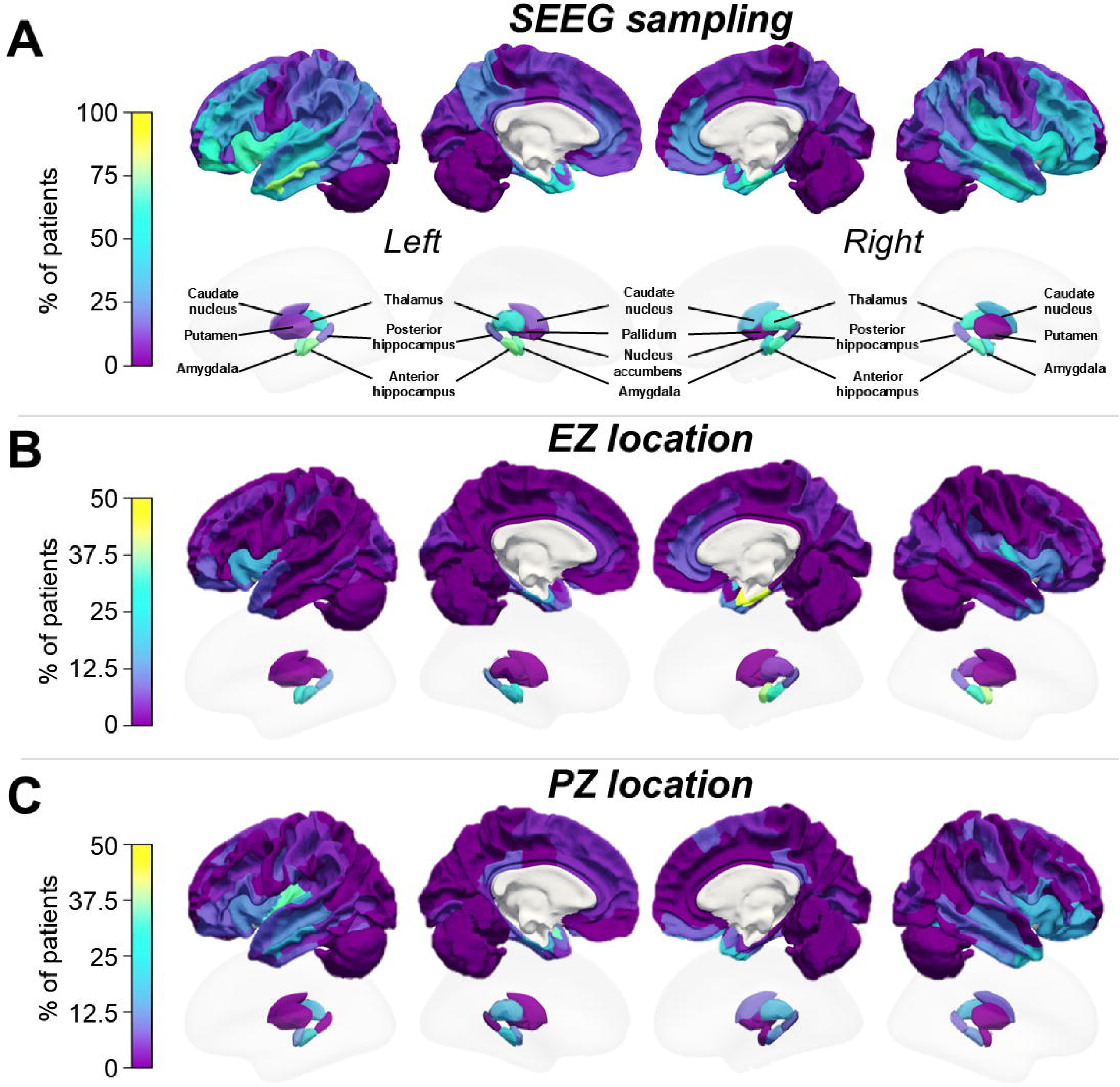
Topography of SEEG sampling, EZ and PZ in the patients cohort. **(A)** For each parcel, percentage of patients for whom at least one SEEG electrode explored the region, based on the VEP atlas parcellation. **(B and C)** For each parcel, percentage of patients for whom that parcel was considered as EZ **(B)** or PZ **(C)**, emphasizing the fact that most of the patients had an involvement of the temporal, orbitofrontal and insular regions in contrast with posterior regions.

**Table 1.** Patients clinical characteristics.

| **Characteristic** | **Overall**<br>**(n = 23)** | **MRI-negative**<br>**(n = 17)** | **Lesional**<br>**(n = 6)** |
| --- | --- | --- | --- |
| Females, n (%) | 12 (52.2) | 6 (35.3) | 6 (100.0) |
| Age, mean [min-max] | 32 [15-56] | 30 [15-53] | 37 [20-56] |
| Handedness, n(%) |  |  |  |
| *Right* | 17 (73.9) | 13 (76.5) | 4 (66.7) |
| *Left* | 5 (21.7) | 3 (17.6) | 2 (33.3) |
| *Ambidextrous* | 1 (4.3) | 1 (5.9) | 0 (0.0) |
| Age at epilepsy onset, mean [min, max] | 17 [0-40] | 16 [0-40] | 19 [8-30] |
| Disease duration (years), mean [min, max] | 15 [6-29] | 14 [6-29] | 19 [6,26] |
| Epileptogenic zone lateralization, n (%) |  |  |  |
| *Left* | 11 (47.8) | 8 (47.1) | 3 (50.0) |
| *Right* | 8 (34.8) | 5 (29.4) | 3 (50.0) |
| *Bilateral R>L* | 4 (17.4) | 4 (23.5) | 0 (0.0) |
| Epileptogenic zone localization, n (%) |  |  |  |
| *Temporal Plus* | 15 (65.2) | 12 (70.6) | 3 (50.0) |
| *Prefrontal Plus* | 4 (17.4) | 3 (17.6) | 1 (16.7) |
| *Posterior* | 2 (8.7) | 1 (5.9) | 1 (16.7) |
| *Insulo-opercular* | 2 (8.7) | 1 (5.9) | 1 (16.7) |
| Radiological diagnosis, n (%) |  |  |  |
| *MRI negative* | 17 (73.9) | 17 (100.0) | 0 (0.0) |
| *Periventricular nodular heterotopia* | 2 (8.7) | 0 (0.0) | 2 (33.3) |
| *Suspected FCD2* | 1 (4.3) | 0 (0.0) | 1 (16.7) |
| *FCD2b* | 1 (4.3) | 0 (0.0) | 1 (16.7) |
| *Ganglioglioma* | 1 (4.3) | 0 (0.0) | 1 (16.7) |
| *Gliosis* | 1 (4.3) | 0 (0.0) | 1 (16.7) |
| Surgery performed, n (%) | 12 (57.1) | 8 (50.0) | 4 (80.0) |
| Surgical outcome, n (%) |  |  |  |
| *Engel I* | 7 (58.3) | 3 (37.5) | 4 (100.0) |
| *Engel II* | 3 (25.0) | 3 (37.5) | 0 (0.0) |
| *Engel III* | 2 (16.7) | 2 (25.0) | 0 (0.0) |
| *Engel IV* | 0 (0.0) | 0 (0.0) | 0 (0.0) |

### Anatomical location of hubs and distribution of sodium parameters

Based on the connectomes averaged across patients and controls, structural connectivity hubs were found in mesial parietal regions, middle and posterior cingulate, as well as in dorso-frontal regions, the insula, the left and right thalami and putamen (Figure 2A). In addition, in patients, the anterior part of hippocampus, orbitofrontal regions, the cerebellum, and caudate nuclei were identified as structural hubs regions as well.

**Figure 2:**
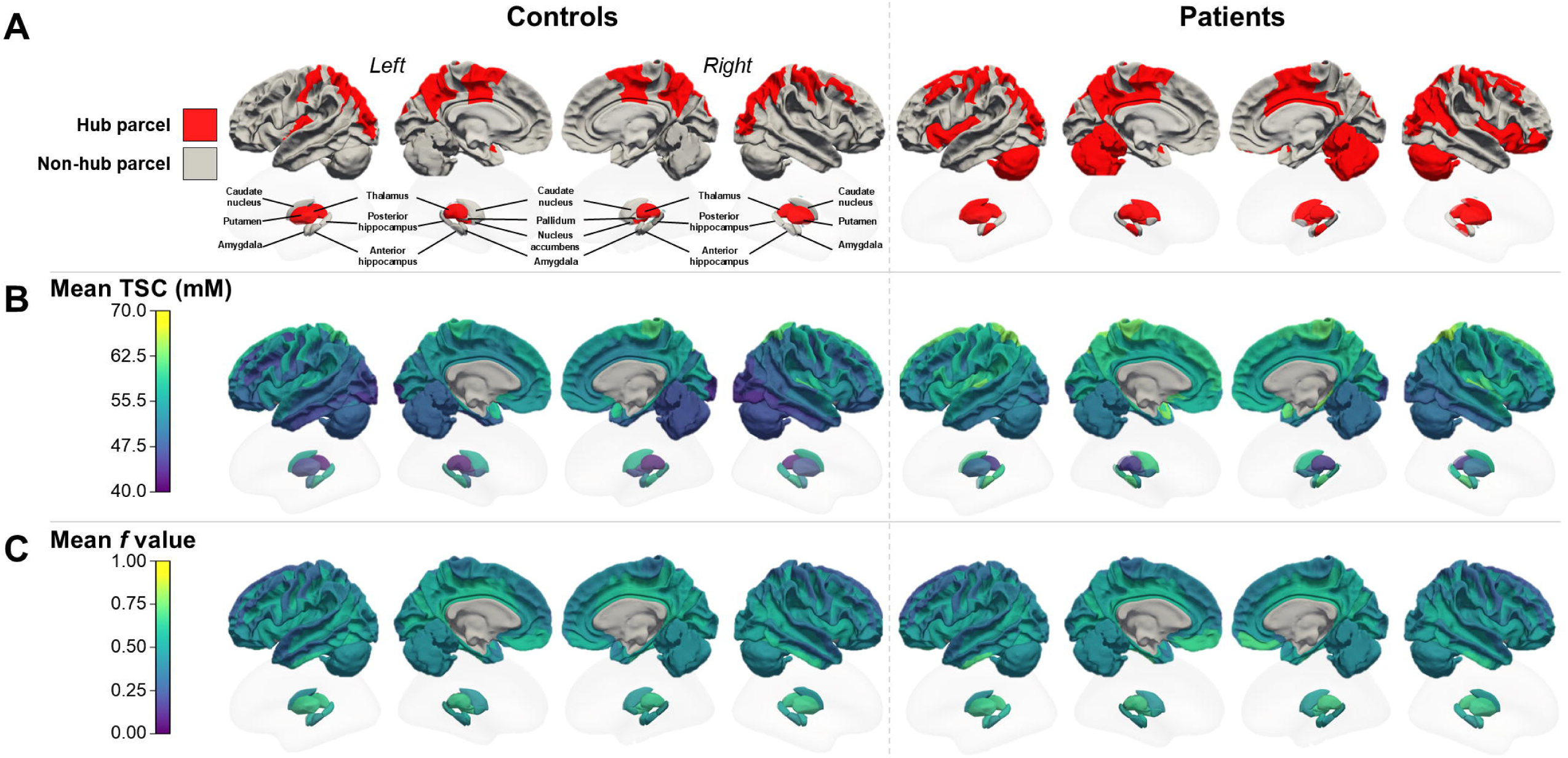
Location of hubs and distribution of TSC and *f* values in the whole cohort. **(A)** Location of hubs parcels of the mean connectome of the controls and the patients respectively. **(B)** mean parcel value of TSC and **(C)** *f*, in the controls and the patients.

At the group level, mean TSC was higher in patients than controls (mean TSC_patients_: 54.6 mM [SD: 7.3]; mean TSC_controls_: 51.3 mM [SD: 7.3], *p* < 10^-4^), and was not homogeneously distributed across brain regions, with increased TSC in midline and peri-insular regions (Figure 2B). In contrast, mean *f* values showed a more homogeneous distribution, with higher mean values found in cingulate and orbito-frontal regions and did not differ between patients and controls (mean *f*_controls_: 0.47 [SD: 0.13]; mean *f*_patients_: 0.47 [SD: 0.14], *p* = 0.1; Figure 2C).

### Effect of epileptogenicity on sodium TSC and *f*

First, we assessed the effect of epileptogenicity on sodium measurements by comparing the TSC and *f* value of each parcel in patients to the median TSC and *f* value of the same parcels in controls. Patients with epilepsy showed an increased TSC in both EZ and non-EZ parcels (Figure 3A, median z-score = +0.51, *p* < 10^-4^). No difference of z-scores distributions was observed when comparing the parcels within the EZ to the parcels within the PZ or NIZ.

**Figure 3:**
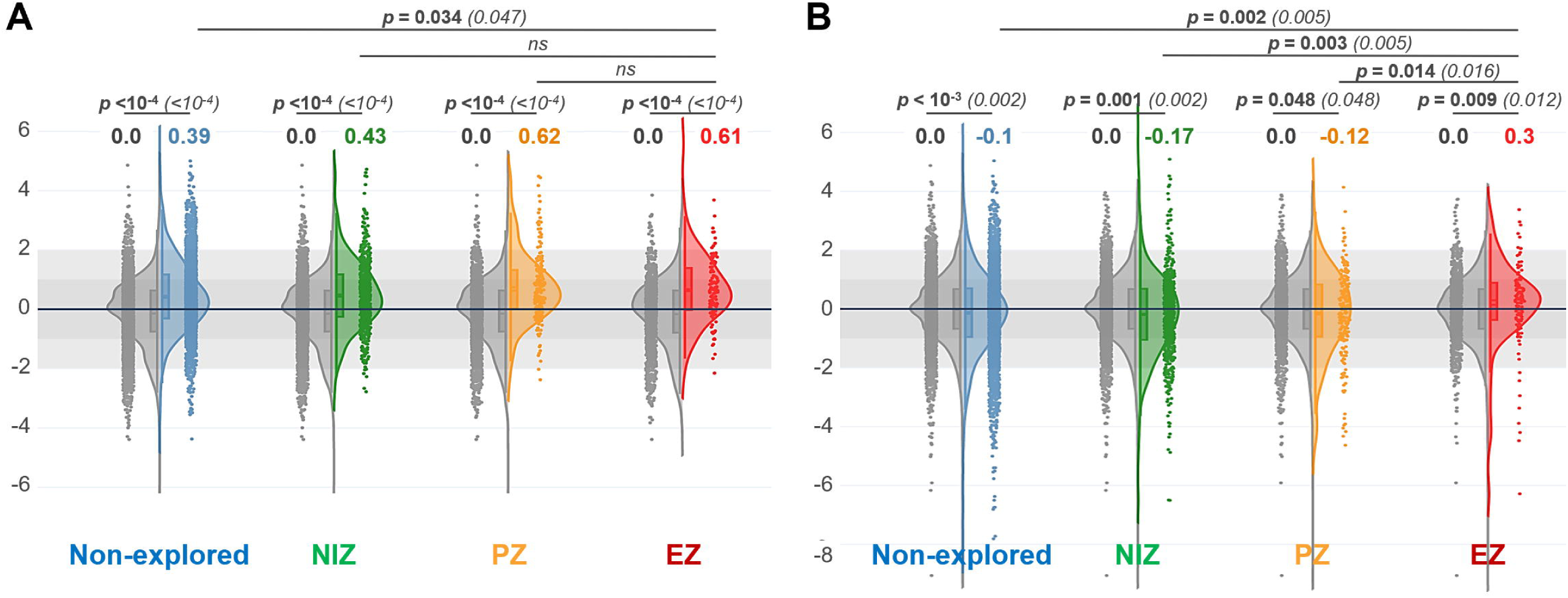
Global increase of TSC while *f* increases only in the EZ. Each patient’s EZ, PZ, NIZ, and non-explored parcels are compared to the homologous parcels of controls. **(A)** Z-score of TSC in patients parcels *vs.* controls. **(B)** Z-score of *f* in patients parcels *vs.* controls.

Contrasting, the *f* value was increased in EZ compared to homologous control parcels (median z-score = +0.3, p = .009, Figure 3B), while in other regions, it was globally lower in epileptic patients compared to controls (median z-scores of -0.17, -0.12, and -0.10 in the NIZ, PZ, and non-explored regions, respectively, p < 0.048).

### Characterization of hub sodium signature

To normalize sodium variations across subjects, in each subject we z-scored each parcel against all parcels of the same subject. We then compared the z-scores of hub parcels with non-hub parcels, in controls and in patients separately (Figure 4A). In controls, hub parcels exhibited increased TSC compared to non-hub parcels (median z-score = +0.21, *p* = .0001, Figure 4B, **Error! Reference source not found.**). The same distribution was observed in patients (median z-score = +0.23, *p* =.0001). On the opposite, no difference was observed in the distributions of the *f* values between hub and non-hub parcels, both in controls and in patients (Figure 4C, **Error! Reference source not found.**).

**Figure 4:**
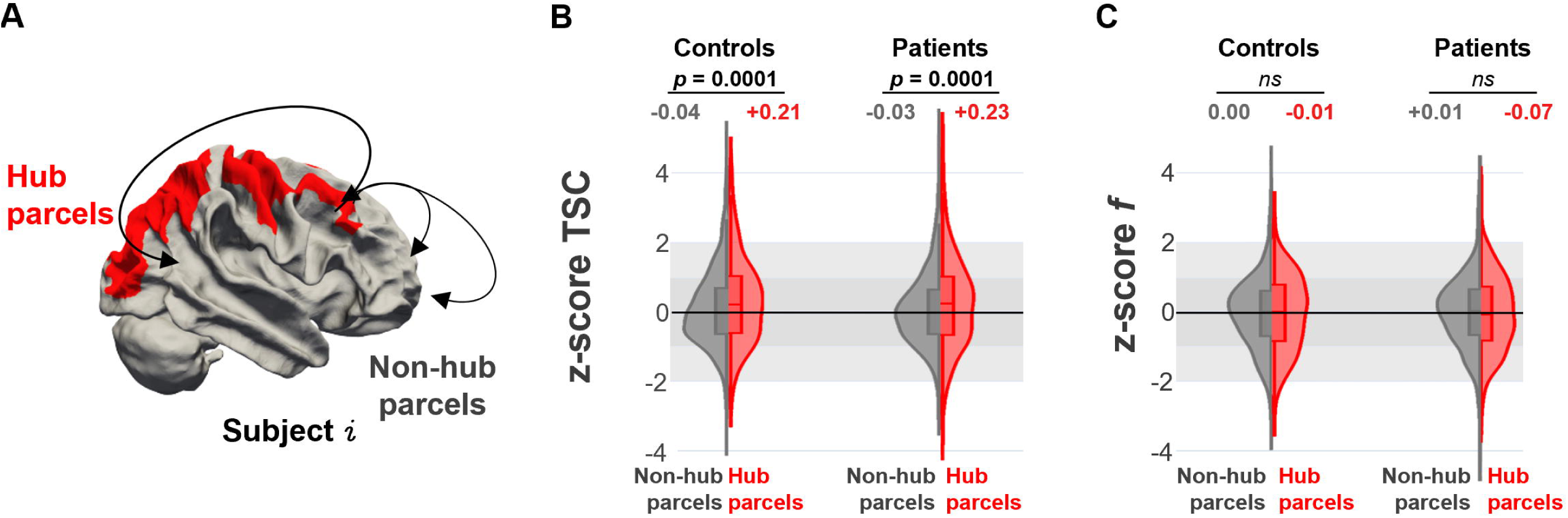
TSC is increased in the hubs of both controls and epileptic patients. **(A)** Each parcel is standardized to the median value of all parcels within each subject, to observe subject-standardized values of **TSC (B)** and *f* **(C)** in controls and patients.

### Influence of epilepsy on sodium homeostasis in hubs

Finally, we assessed the interaction of epilepsy and hubness on sodium measures in patients compared to controls. We compared the TSC and *f* values of the hub and non-hub parcels of patients to the median TSC and f values of the same parcels in all controls (Figure 5A). An increase of TSC was observed in parcels of patients compared to controls, regardless of whether the parcel was a hub (median z-score = +0.34, *p* < 10^-4^) or not (median z-score = +0.43, *p* < 10^-4^, Figure 5B). Again, we observed a slight decrease of the *f* value in patients compared to controls, but this was not dependent on the presence of a hub (median z-score = -0.13, *p* = .002) or not (median z-score = -0.09, *p* < 10^-4^, figure 5C).

**Figure 5:**
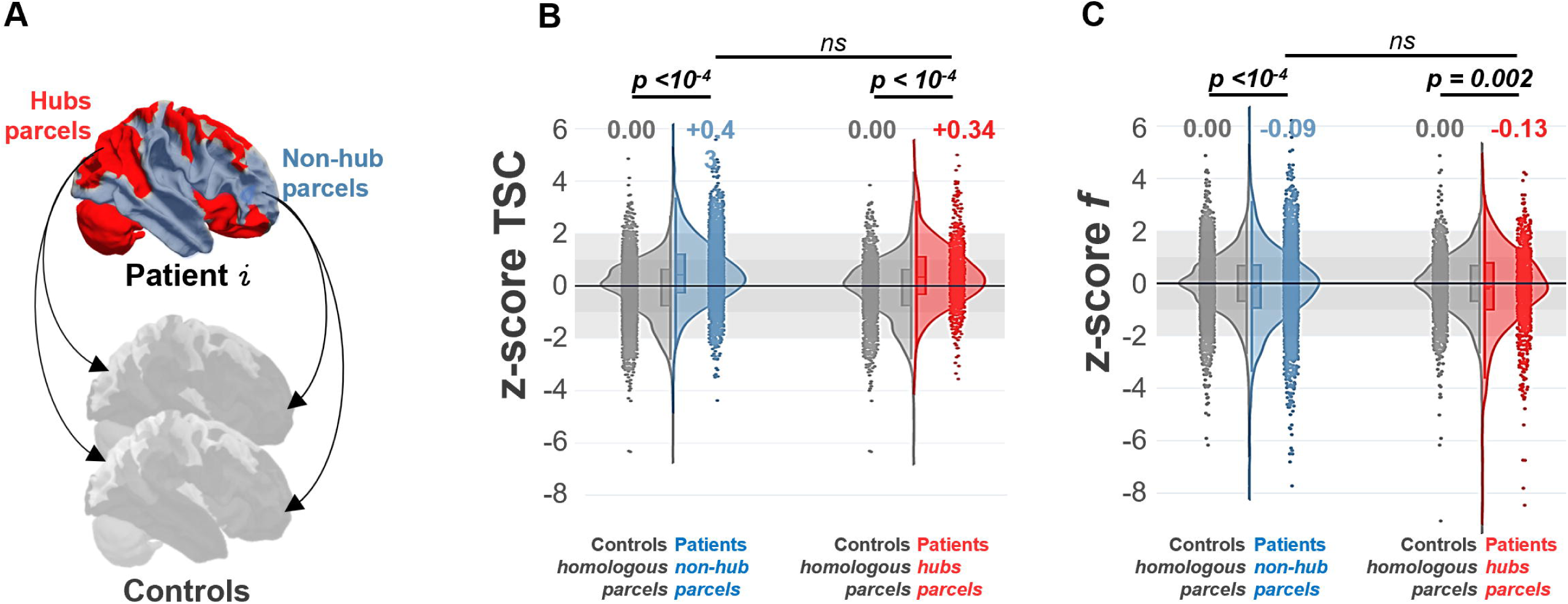
The increase of TSC in the hubs is seen both in controls and patients. **(A)** Each patient’s parcels values are standardized to the median value of homologous parcels in controls. This allows a patient *vs.* controls comparisons based on the z-score of TSC **(B)** and *f* **(C)** in hubs and non-hubs parcels.

### When structural hubs are part of the epileptogenic network

We assessed if sodium measures could be different in patients with at least one hub parcel overlapping with the EZ parcels (10/23 patients), and those without any hub involved in the EZ (13/23 patients). For each group, we compared parcel’s TSC and *f* to the same parcels in controls (Figure 6).

**Figure 6:**
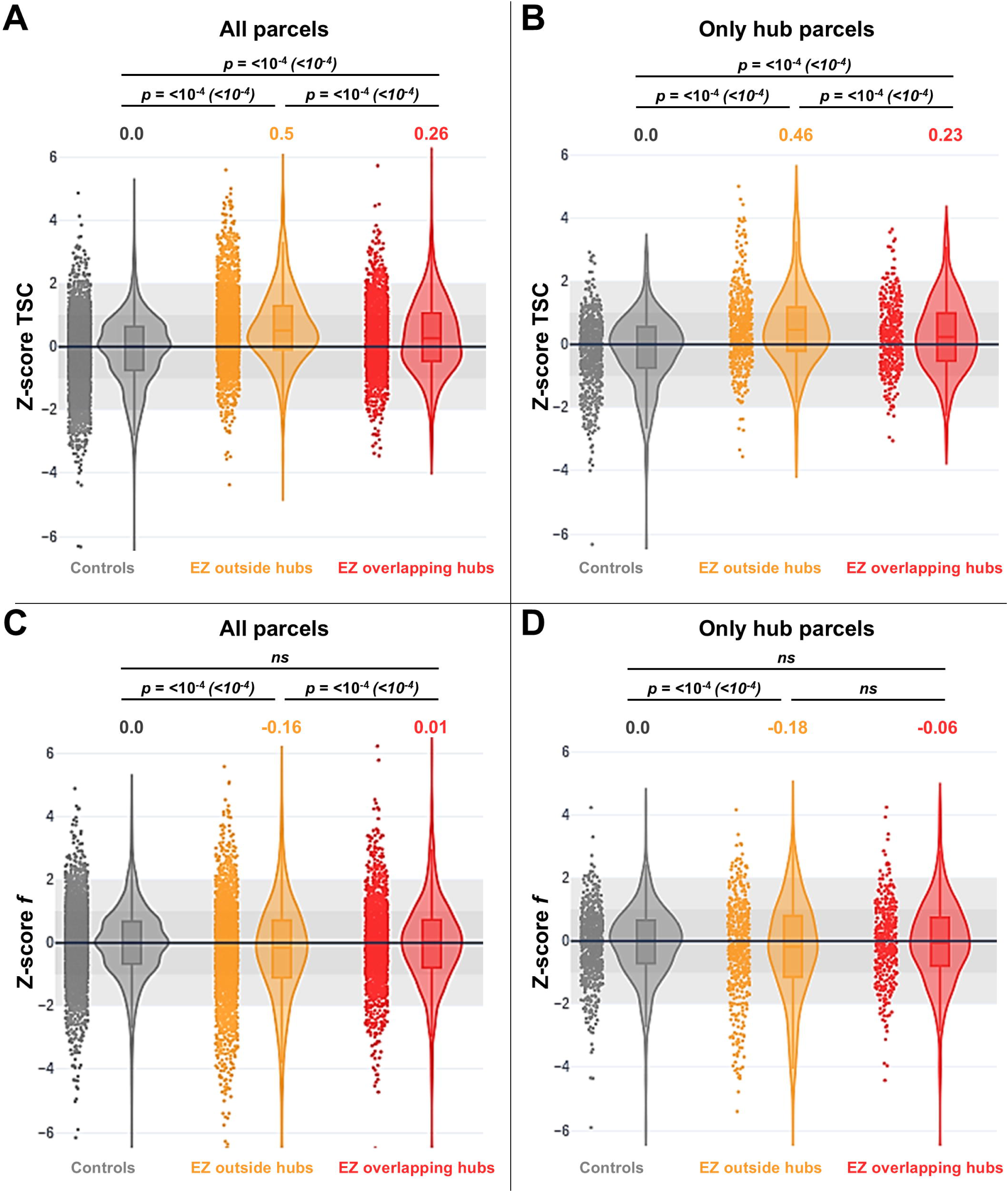
Differences in TSC and *f* when hubs and EZ overlaps. First row **(A, B)** shows TSC values, second row **(C, D)** shows *f* values. Each patient’s parcels values are standardized to the median value of homologous parcels in controls. **(A)** and **(C)** show TSC and *f* respectively for all parcels of patients, whether hubs and EZ overlap or not, compared to controls. **(B)** and **(C)** show TSC and *f* respectively only in hubs in patients, compared to controls.

The increase of TSC and decrease of *f* in parcels of patients we described above was more pronounced in patients without overlap between the EZ and hub parcels (median z-score TSC: +0.5 vs. +0.26*, p* < 10^-4^; median z-score *f*: -0.18 vs. -0.06*, not statistically significant)*. However, we did not observe statistically significant differences in the extent of the EZ in both groups: patients (Patients without EZ overlapping hubs have in average 4 EZ parcels *vs.* 4.5 parcels in patients with EZ overlapping hubs, or, in the clinical features of both groups (Supplementary Table 1). In patients, we observed a positive correlation between the median TSC and the extent of the EZ network (defined by the number or EZ or EZ+PZ parcels), but not with the median *f*; and there was no statistical difference of this correlation between patients with or without EZ and hub overlap (Supplementary Fig. 4).

## Discussion

Our study aimed to investigate the characteristics of sodium homeostasis within structural connectivity hubs using ultra-high field ^23^Na MRI, and to determine how it may be affected in DRFE.

Our results confirm the diffuse increase in TSC observed in the cortex in epileptic rats, and later in patients with DRFE at 3T and then 7T, which appeared unspecific to the epileptogenicity of a specific region.^55,21,22^ Moreover, we observed that the global increase of TSC is associated with the extension of the EZ network. These observation aligns with the increases in TSC observed across several brain diseases.^18,20,56–59^ Only one study reported a consistent decrease of TSC in patients with mild traumatic brain injury, and one study reported a decrease of TSC in the case of one patient having a seizure during the scan.^21,60^

Furthermore, using 7T MRI and multiple echoes ranging from 0.2 ms to 70.78 ms allows for measuring the sodium signal decay and extracting the parameters of this decay (here, the *f* ratio). Recently, our group observed an increase in the *f* ratio in spherical regions of interest centered around depth electrodes contacts located within the EZ, in contrast to electrodes positioned in the PZ or the NIZ.^22^ We now replicated these findings at the parcel level and in a bigger patient cohort (12/23 patients and 5/21 controls shared with the previous cohort), demonstrating an increase in the *f* ratio within the EZ of the patients. This increase was evident when comparing the EZ parcels to the same parcels in controls and non-EZ parcels in patients. Moreover, we observed a decrease in the *f* ratio in patients’ non-EZ parcels compared to the homologous parcels in the controls.

### Sensitivity of ^23^Na signal to tissular integrity and epileptogenicity

The diffuse increase of TSC in the context of focal epilepsy does seem to correlate with brain-wide pathological alterations, but not with the epileptogenicity of a particular region. Indeed, focal epilepsies are associated with local alterations in brain microstructure as demonstrated by histological techniques for several decades, even in the absence of macroscopic lesions.^61–63^ Moreover, studies have shown that microstructural changes are not limited to the EZ, but also occur in anatomically distant regions.^64–67^ These studies are consistent with the observation of diffuse cognitive alterations that cannot solely be explained by pathological processes limited to the networks involved in the EZ.^68^ Therefore, TSC may be an additional biomarker of these diffuse microstructural alterations in the context of focal epilepsies.

On the opposite, the variations in sodium signal fraction (*f*) we observed in the EZ seem to be related to the epileptogenic characteristics of a region, independently of TSC. While *f* reflects the degree of the tissue’s cellular organization, providing insights into intracellular compartments, the physiological underpinning of these variations remains unclear. In one study investigating the correlation between the sodium signal and tissue microstructure using diffusion MRI, no clear relationship was found between the fast component of sodium decay and white or grey matter densities.^69^ The authors have suggested that regional differences in magnetic susceptibility may explain some of the variations, but this has yet to be fully evaluated. To our knowledge, aside from this study and the previous study of our group (Azilinon et al. 2022^22^), no other published studies examined sodium signal fraction in the context of neurological conditions.

### Total sodium concentration differs between hub and non-hub regions

Interestingly, our study revealed that TSC was elevated in hub regions compared to non-hub regions, in both controls and patients, while *f* remained unchanged. From the physiological point of view, structural hubs are regions of high metabolic cost (conceptualized as a “wiring cost”, and measured with PET studies), and are often found at the epicenter of neurological diseases.^27,70^ In the context of focal epilepsy, structural hubs are particularly prone to atrophy.^71^ This highlights the microstructural specificities of hubs that may be reflected with increased TSC compared to non-hub regions.

Also, these hub cortical regions are thinner than other parts of the cortex.^72^ As a consequence, some contribution of CSF partial volume effect could partially contribute to the observed increase in TSC, even though we were careful to exclude voxels containing more than 10 % CSF probability using high-resolution segmentation and restrictive nearest neighbor interpolation when reslicing in sodium space. Another limitation of this work is the consistency of the major structural hub regions between subjects (the dorso-frontal cortex, the cingulum, the precuneus and the thalamus).^73,27^ Because “non-hub precuneus” or “non-hub thalami,” for instance, do not exist as references, the “hubness” of a region cannot be studied in isolation from its anatomical location, and it is not possible to examine the isolated effect of “hubness” on the sodium signal. Since these regions have histological characteristics that differ from other brain parts (e.g., cortical thickness, cell density, neuron/glial cell ratio, myelin content)^74^, the increase of TSC in these regions may also reflect related microstructural differences. In our opinion, choosing an arbitrary set of regression variables to try to correct for regional effects would be difficult to interpret given these many and intricate parameters and the size of the cohorts we are studying.

### Linking structural connectivity, sodium homeostasis and epileptogenicity

Contrary to our initial hypothesis, we did not observe epilepsy-related variations in sodium signal fraction between the different types of structural connectivity nodes, *i.e.* hubs versus non-hubs. The *f* ratio did not follow the regional differences of TSC observed between hub and non-hub regions in either controls or patients, suggesting that sodium signal fraction is less affected by the local microstructure than TSC.

Interestingly, the canonical hubs (*i.e.,* the precuneus, thalamus, and dorso-frontal region) were rarely involved in the EZ network in our patient cohort (Figure 2 and 3). First, our cohort excludes patients with self-limited or drug-responsive focal epilepsy (like self-limited epilepsy with centrotemporal spikes or childhood occipital visual epilepsy), and it does not include patients with generalized epileptic syndromes. The underlying physiology of these syndromes is very different from that of DRFE and conclusions drawn on DRFE may not apply to these other forms of epilepsy.^75^ Second, aside from surgical considerations (i.e. the difficulties of resecting highly functional regions), the EZ of DRFE most commonly involves the temporal regions (with temporal-frontal, temporal-insular, or temporal-parietal networks) followed by the frontal regions (frontal-orbital, anterior cingulate, or dorsal frontal regions) and, more rarely, the occipital and parietal regions.^76–78^ Several hypotheses have been proposed to explain the predisposition of the temporal lobe to be the epicenter of DRFE, either based on observations of a specific expression of excitatory neurons, or a vulnerability to develop epileptic activities based on the neural circuits specialized in memory and emotion processing compared to circuits specialized in visuo-spatial processing.^79–81^

Furthermore, there is growing evidence supporting the concept of “isolation” of the EZ in focal epilepsy. Contrasting with the idea of the EZ acting as a pathological hub at the scale of the whole brain, several studies suggest that the EZ exhibits strong functional or structural coupling within its own network, but decreased connectivity with regions outside the EZ.^82–86^ While the term “isolation” could be misleading—since it suggests the EZ is completely disconnected from the rest of the brain—connectivity measurements and graph metrics offer a different perspective. They support the hypothesis that the EZ of focal epilepsies consists of a network of strongly interconnected regions with reduced control from other cortical regions, making these “peripheral autonomous regions” more prone to homeostatic dysregulations and to develop the capacity to generate autonomous activities.^87,88^ Based on the observation of a loss of correlation between the *f* value and energy metabolism markers within the EZ, as opposed to the PZ or the NIZ,^22^ we hypothesize that the sodium signal fraction f could potentially be sensitive to the degree of autonomy of the EZ.

### Networks and hubs are not involved equally in every DRFE patient

Previous studies have suggested that the involvement of hubs in the epileptogenic network is associated with a higher risk of seizure recurrence after epilepsy surgery.^31,89^ While our study did not reach statistical significance, we observed a trend in which patients with MRI-negative epilepsy and seizure recurrence after surgery were more likely to have parts of the EZ overlapping with structural connectivity hubs. In contrast, patients with mono-focal lesion-related epilepsy (such as FCD2b or ganglioglioma) did not had hub-EZ overlap and more often achieved better surgical outcomes, classified as Engel class I. Although these observations require further confirmation, they are consistent with previous studies and suggests two key considerations. First, we may need to distinguish between epilepsies driven by a monofocal, anatomically well-delineated epileptogenic lesion (and whether or not it is located within a structural hub) and “MRI-negative, histology-negative epilepsies with focal seizures” which may involve a broader disruption of the functional network of unknown origin. Second, it is crucial to differentiate between “structural” and “functional” hubs (as identified through EEG, MEG, or fMRI) and to clarify the scale at which these hubs are considered—whether at the whole-brain, lobar, or sublobar level.

## Conclusion

Our findings indicate that the observed increase of TSC in structural hub regions is more likely related to microstructural abnormalities and general unspecific loss of tissular integrity rather than specific to epilepsy. We also confirmed the increase in the *f* value in the EZ observed in previous studies. This increase may serve as a marker of susceptibility to homeostasis failure associated with the generation of abnormal rhythms, a phenomenon not seen in the structural hubs of DRFE patients. We propose that sodium signal fraction may be more closely linked to the homeostasis of a region than its microstructure. Future research could focus on exploring the correlation between sodium relaxometry, functional connectivity and gene expression maps, and examine how the dynamics of the epileptogenic network in DRFE could be further elucidated through sodium imaging.

## Supporting information

STROBE

## Acknowledgements

This work was performed by a laboratory member of France Life Imaging network (grant ANR-11-INBS-0006).

## Supplementary material

Supplementary material is available at *Brain* online.

