## Supplementary for "Investigating sodium homeostasis of structural brain hubs in focal epilepsy using 7T MRI"

### **Supplementary Figure** **1**

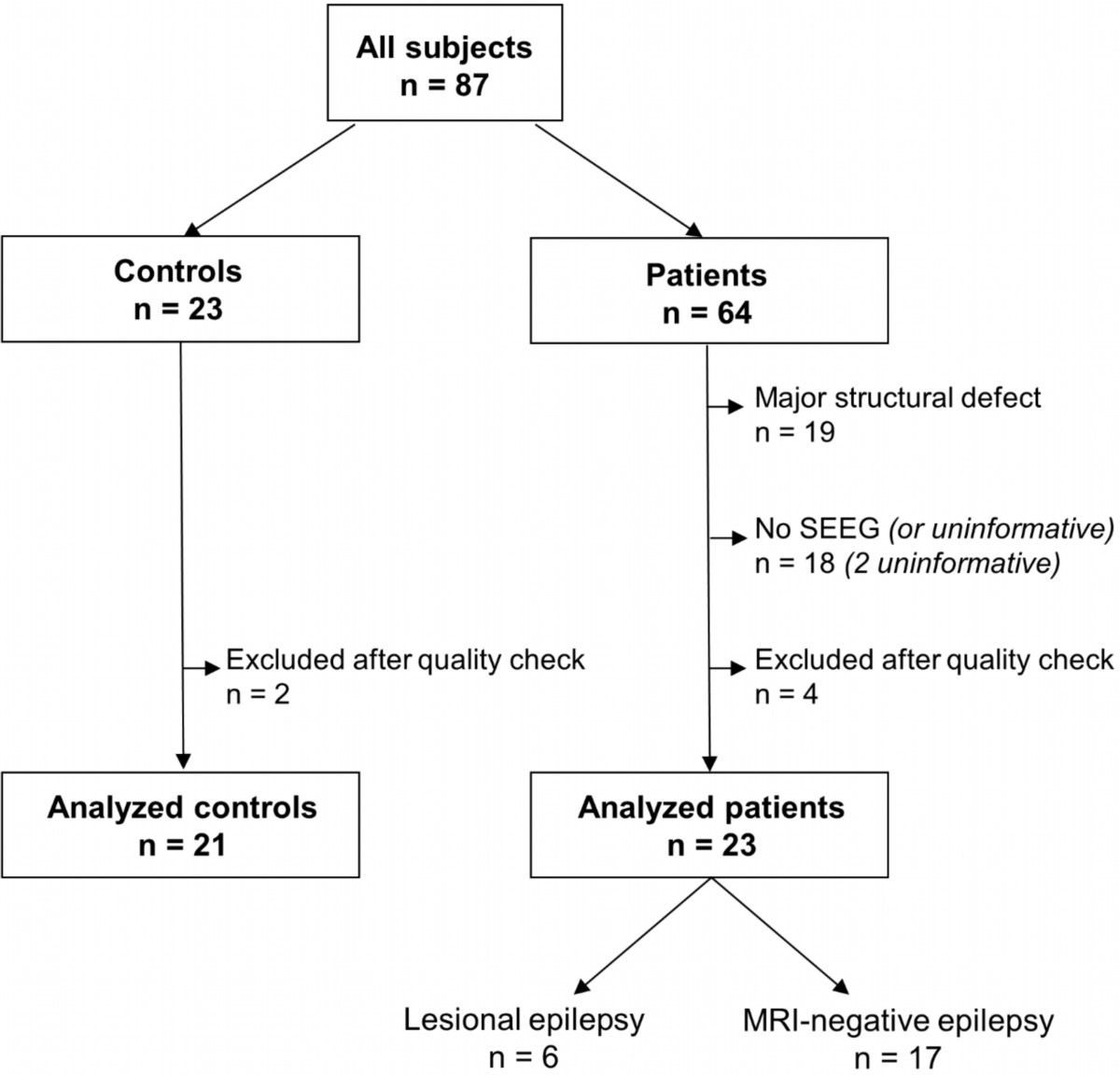

**Supplementary Figure 1: Flow chart of the selection of controls and patients.**

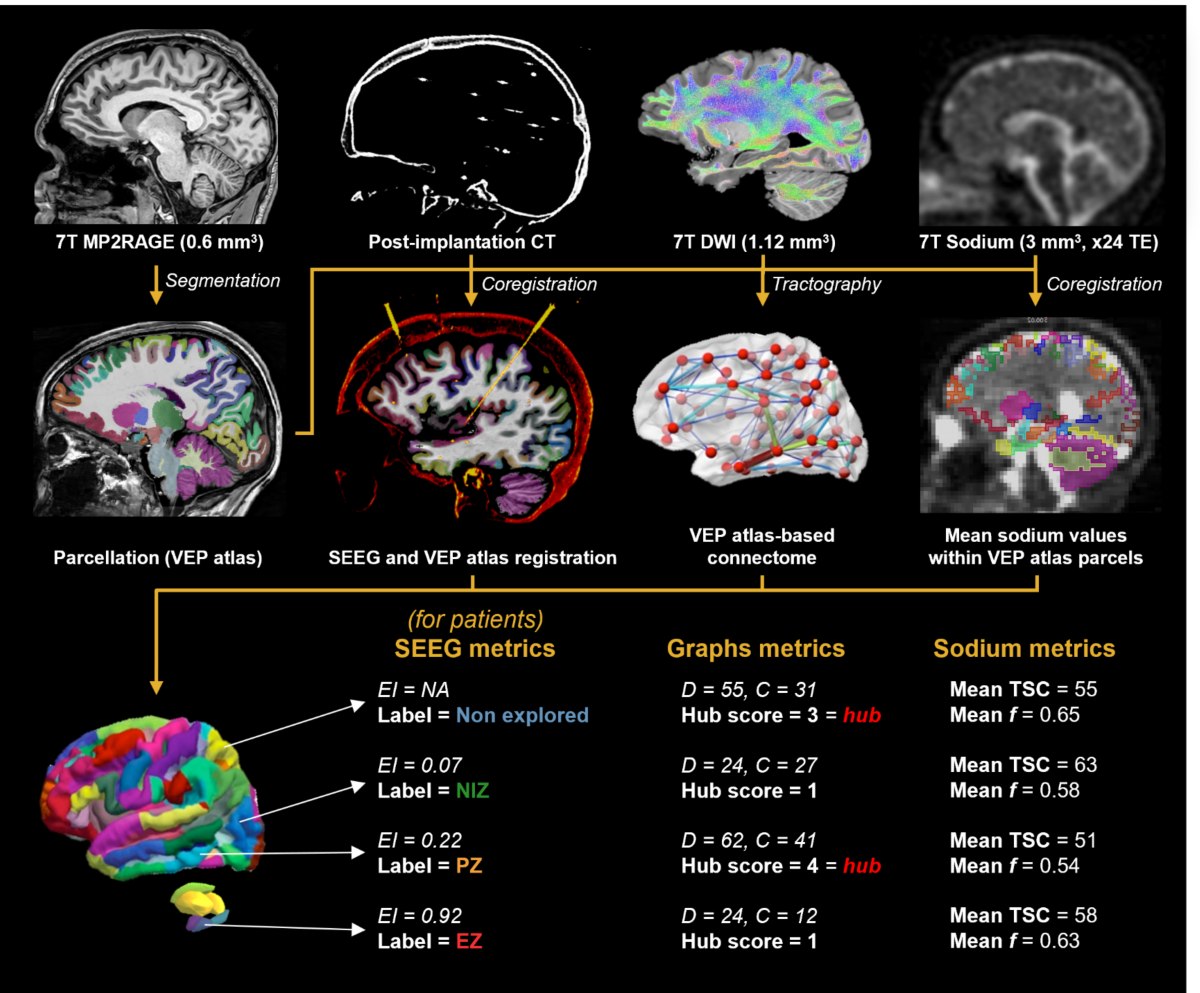

### **Supplementary Figure 2**

**Supplementary figure 2: Graphical summary of the processing pipeline**. Segmentation, parcellation and post-implantation CT coregistration are performed on high-resolution MP2RAGE sequences to define parcels and their contribution to the epileptogenic network based on SEEG exploration. Hubs are defined based on the connectomes derived from DWI sequences and anatomical parcellation. Finally, the anatomical parcellation is resliced in sodium space to extract the mean TSC and *f* of each region.

### Supplementary File 1

#### DWI Tractography processing parameters

##### Tract generation (MrTrix3):

tckgen ‑act 5TT.nii.gz ‑crop_at_gmwmi ‑seed_gmwmi gray_white_interface.nii.gz ‑algorithm iFOD2 ‑select 10M ‑minlength 20 ‑nthreads 6 CSD_wm_norm.nii.gz 10M_act_tracks.tck

Options used:

- -algorithm iFOD2: Second-order Integration over Fiber Orientation Distributions.
- -act: use the Anatomically-Constrained Tractography framework during tracking.
- -select 10M: desired number of streamlines to be selected: 10 million here.
- -seed_gmwmi: seed from the grey matter - white matter
- -crop_at_gmwmi: crop streamline endpoints more precisely as they cross the GM-WM interface.
- -minlength 20: minimum length of any track in mm.

*Filtering (SIFT algorithm available in MRtrix3)*

tcksift 10M_act_tracks.tck CSD_wm_norm.nii.gz 10M_act_tracks_sift.tck ‑term_number 1000000 ‑force ‑act 5TT.nii.gz

Options used:

- -term_number 1000000: filtering to 1 million fibers.
- -act: ACT five-tissue-type segmented anatomical image to derive the processing mask.

*Connectome generation (MRtrix3)*

labelconvert aparc+aseg_vep_to_diff.nii.gz VepFreeSurferColorLut.txt labels_aparc_aseg_Vep.txt parcellated.nii.gz

tck2connectome ‑zero_diagonal ‑symmetric 10M_act_tracks_sift.tck parcellated.nii.gz VEP_connectome_N.csv ‑out_assignment assignments_N.csv

Options used:

- ‑zero_diagonal
- -symmetric

### S**upplementary Figure 3**

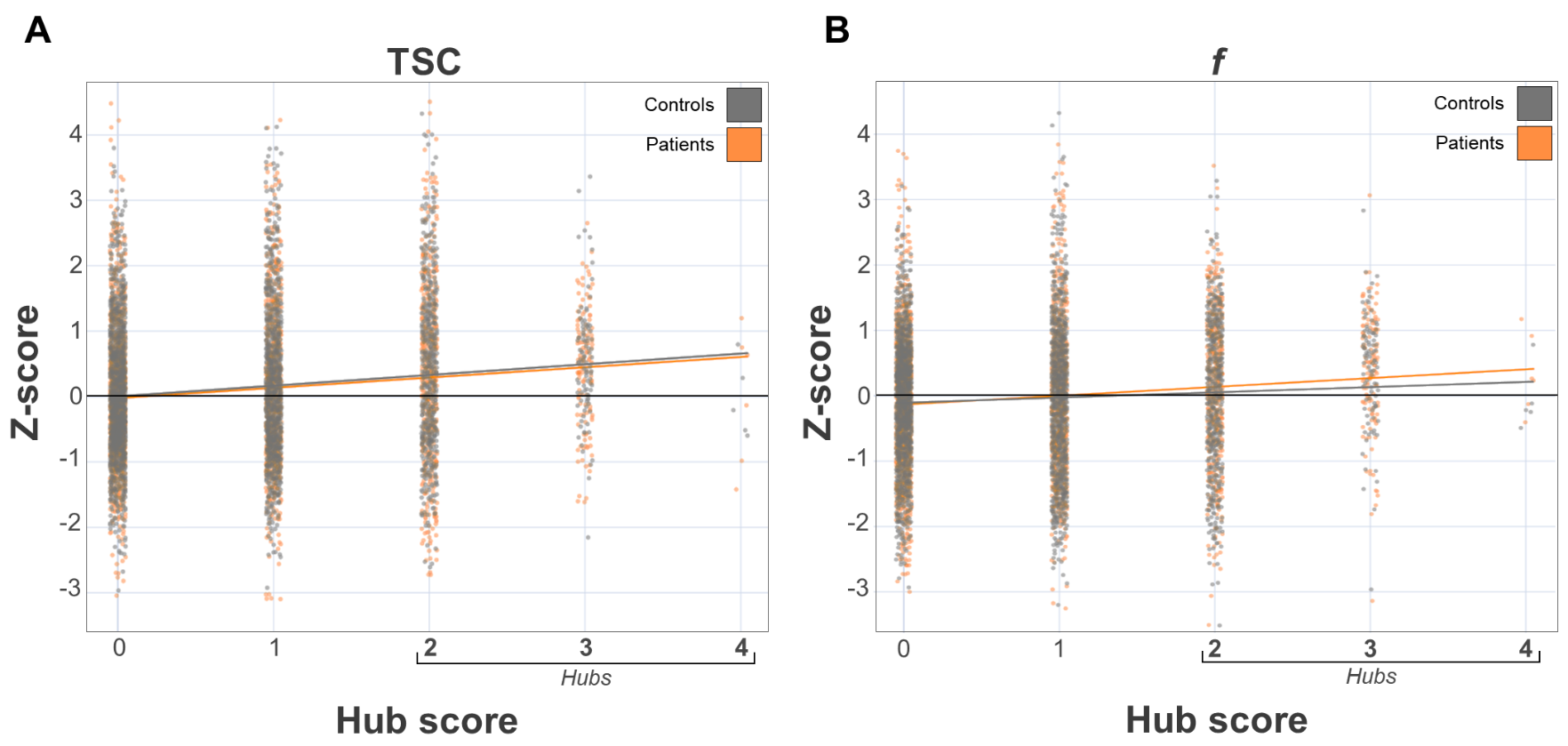

Supplementary Figure 3 : Relation between parcels hub score and sodium signal. For each subject, each parcel TSC (A) or *f* (B) value is z-scored against all parcels of the subject.

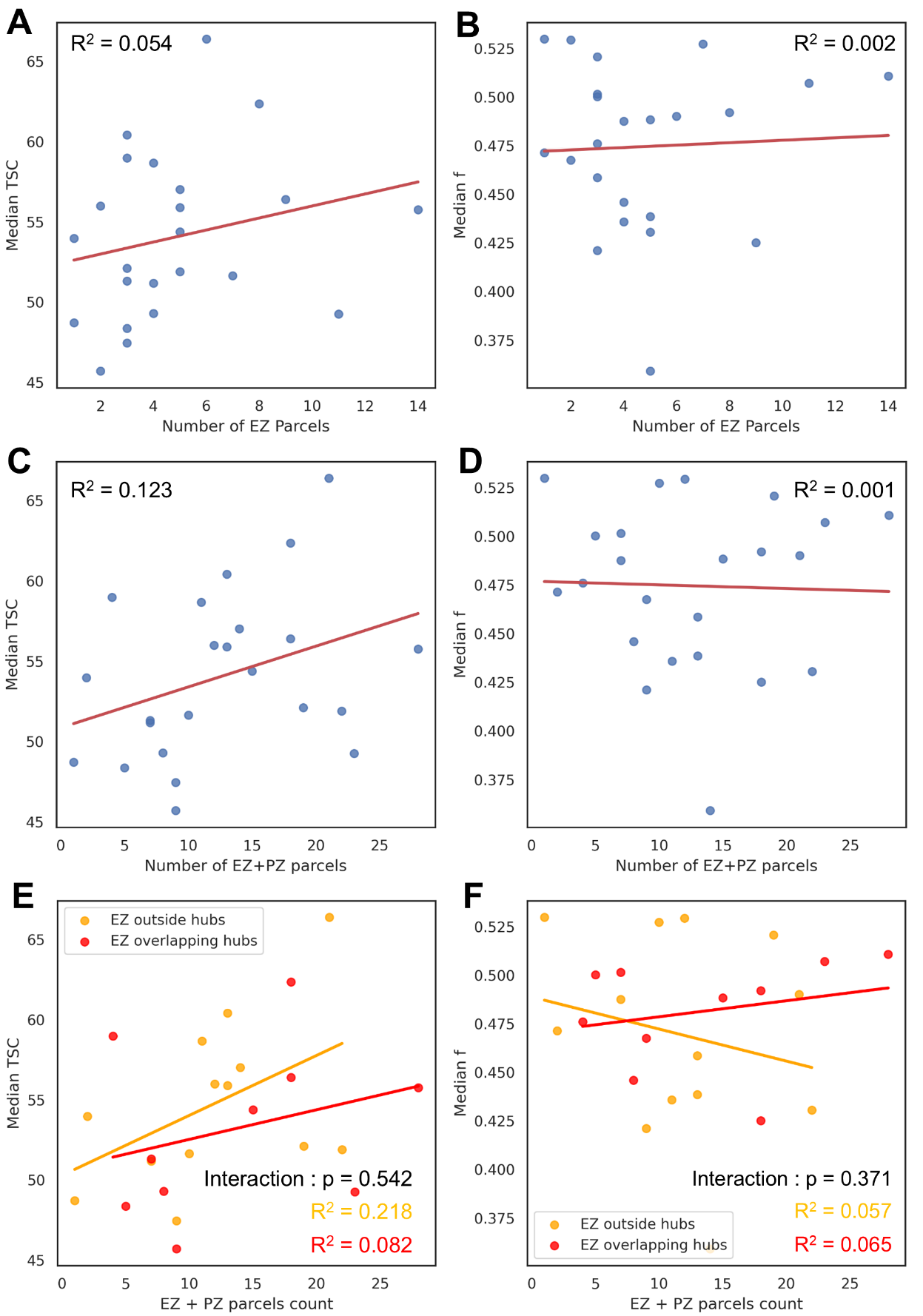

### Supplementary Figure 4

**Supplementary Figure 4**: Relation between median TSC or f and extension of the EZ or PZ. Median TSC **(A)** or *f* **(B)** compared to number of EZ parcels in patients. Median TSC **(C)** or *f* **(D)** compared to number of EZ and PZ parcels in patients. Median TSC **(E)** or *f* **(F)** compared to number of EZ and PZ parcels in patients with hubs overlapping the EZ network or not.

### S**upplementary Table 1**

**Supplementary Table 1 Clinical characteristics and outcomes when EZ overlap with hubs**

|  | **Missing** | **No EZ/Hub overlap**  **(*n* = 13)** | **EZ/Hub overlap**  **(*n* = 10)** | ***p*-value** |
| --- | --- | --- | --- | --- |
| Females, n (%) | 0 | 7 (53.8) | 5 (50.0) | 1.000 |
| Age, mean [min-max] | 0 | 34 [15-56] | 28 [16-53] | 0.154 |
| Handedness, n(%) | 0 |  |  | 0.285 |
| *Right* |  | 8 (61.5) | 9 (90) |  |
| *Left* |  | 4 (30.8) | 1 (10.0) |  |
| *Ambidextrous* |  | 1 (7.7) | 0 (0.0) |  |
| Age at epilepsy onset, mean [min, max] | 0 | 12 [2-40] | 16 [0-40] | 0.756 |
| Disease duration (years), mean [min, max] | 0 | 14 [8-29] | 10 [6,29] | 0.121 |
| Epileptogenic zone lateralization, n (%) | 0 |  |  | 0.231 |
| *Left* |  | 8 (61.5) | 3 (30.0) |  |
| *Right* |  | 4 (30.8) | 4 (40.0) |  |
| *Bilateral R>L* |  | 4 (7.7) | 3 (30.0) |  |
| Epileptogenic zone localization, n (%) | 0 |  |  | 0.343 |
| *Temporal Plus* |  | 10 (76.9) | 5 (50.0) |  |
| *Prefrontal Plus* |  | 2 (15.4) | 2 (20.0) |  |
| *Posterior* |  | 1 (7.7) | 1 (10.0) |  |
| *Insulo-opercular* |  | 0 (0.0) | 2 (20.0) |  |
| Lesion-related epilepsy, n (%) | 0 | 4 (30.8) | 2 (20.0) | 0.660 |
| Radiological diagnosis, n (%) | 0 |  |  | 0.330 |
| *MRI negative* |  | 9 (69.2) | 8 (80.0) |  |
| *Periventricular nodular heterotopia* |  | 0 (0.0) | 2 (20.0) |  |
| *Suspected FCD2* |  | 1 (7.7) | 0 (0.0) |  |
| *FCD2b* |  | 1 (7.7) | 0 (0.0) |  |
| *Ganglioglioma* |  | 1 (7.7) | 0 (0.0) |  |
| *Gliosis* |  | 1 (7.7) | 0 (0.0) |  |
| Surgery performed, n (%) | 2 | 8 (61.5) | 4 (50.0) | 0.673 |
| Surgical outcome, n (%) | 0 |  |  | 0.062 |
| *Engel I* |  | 5 (62.5) | 2 (50.0) |  |
| *Engel II* |  | 3 (37.5) | 0 (0.0) |  |
| *Engel III* |  | 0 (0.0) | 2 (50.0) |  |
| *Engel IV* |  | 0 (0.0) | 0 (0.0) |  |
| WAIS IV: IQT, median [min-max] | 4 | 80 [70-101] | 75.5 [56-109] | 0.254 |
